# Impact of Connecticut’s 2021 Repeal of Religious Vaccine Exemptions on Kindergarten Vaccine Coverage

**DOI:** 10.64898/2026.06.19.26356105

**Authors:** Casey Benzaken, Julia Moniz Ganem, Barbara L. Araujo, Camila Aparicio-Llorente, Isabela O. Oliva, Aanchal Wats, Diego R Hijano, Carlos Oliveira

## Abstract

**Importance:** Religious vaccine exemptions remain central to debates over school-entry immunization mandates, but evidence on exemption repeal outside outbreak-driven policy responses and across communities with different religious contexts remains limited.

**Objective:** To estimate changes in kindergarten vaccination coverage associated with Connecticut’s 2021 repeal of religious vaccine exemptions, examine variation by school type and local religious congregation density, and compare trends with states that retained exemptions.

**Design/Setting/Participants:** Interrupted time series analysis using kindergarten vaccination data from 2012-2025. Vaccine coverage trends in Connecticut were compared to Arizona, Louisiana, and Oregon, which retained religious exemptions during the study period.

**Intervention:** The intervention studied was Public Act 21-6, which eliminated religious vaccine exemptions from school-entry immunization requirements.

**Main Outcomes and Measures:** Outcomes included annual coverage for measles-mumps-rubella (MMR), varicella, diphtheria-tetanus-acellular pertussis (DTaP), polio, and hepatitis B vaccines. Models estimated pre-policy trends, immediate level changes, and post-policy slope changes. Analyses were stratified by public and private schools and by county-level religious congregation density.

**Results:** Before policy implementation, kindergarten vaccination coverage in Connecticut declined across all vaccines by 0.16-0.20% per year (p < 0.001). Repeal of the religious exemptions occurred during a period of increasing religious congregation density and was associated with improved school-entry vaccination coverage, with annual coverage increasing 0.88-1.02% per year (p < 0.001). Coverage increased in both public and private schools, with larger post-policy gains in private schools. Coverage increases did not differ significantly between high- and low-religiosity counties. In segmented regression analyses, Connecticut’s post-policy MMR slope was significantly higher than those of Arizona, Oregon, and Louisiana by 1.36, 1.71, and 1.15 percentage points per year, respectively (p < 0.001). By 2024/25, Connecticut MMR coverage reached 98.2%, exceeding coverage in comparison states by 5.6-9.6%. Cumulatively, the model-estimated policy impact represented an estimated 2,579 additional kindergarteners immunized against MMR compared with the no-policy counterfactual.

**Conclusions and Relevance:** Connecticut’s repeal of religious vaccine exemptions was associated with increases in kindergarten vaccination coverage across public and private schools, independent of local religious congregation density. These findings suggest that removal of religious vaccine exemptions may be an effective policy approach to improve childhood immunization coverage.

**KEY POINTS:** *Question:* How does Connecticut’s repeal of religious vaccine exemptions affect kindergarten vaccination coverage in the context of local religious and nationwide coverage trends?

*Findings:* An interrupted time series analysis using data from 2012-2025 revealed that repeal of religious vaccine exemptions was followed by a reversal of declining kindergarten vaccination coverage trends. Coverage increases did not differ significantly between high- and low-religiosity counties. Compared with states that retained religious exemptions, Connecticut had significantly greater post-policy increases in vaccination coverage.

*Meaning:* Repeal of religious vaccine exemptions may improve school-entry vaccination coverage even during a period of increasing religious congregation density and may reduce disparities between public and private schools.

## Introduction

Vaccine mandates have long played a critical role in preventing outbreaks of vaccine-preventable diseases, especially in school-aged populations.^1–4^ While all U.S. states allow medical exemptions to these requirements, many have also allowed nonmedical exemptions, including those based on religious or philosophical beliefs.^5–7^ Religious exemptions in particular have become a major source of policy debate because they can contribute to geographic clustering of under-immunized children, which has facilitated outbreaks even when statewide coverage remains high.^4,8–10^ Recent studies further suggest that religious exemptions may not always reflect formal doctrinal opposition to vaccination but rather function as a substitute pathway for broader vaccine refusal from secular concerns or personal skepticism.^11–14^

These concerns have prompted several states to enact legislation to restrict or eliminate religious and nonmedical exemptions. California, for example, eliminated all nonmedical exemptions in 2016, and New York State eliminated religious exemptions in 2019.^15,16^ Studies evaluating these policy changes have shown significant increases in school-entry vaccination coverage following implementation.^17–20^ However, because many of these policy changes occurred during outbreaks or periods of heightened public concern about vaccine-preventable disease, it remains unclear whether similar effects would be observed in a non-crisis context.

In April 2021, Connecticut passed Public Act 21-6 (PA 21-6), making it one of six U.S. states to eliminate religious exemptions for school-required vaccinations, retaining only medical exemptions. This rule became effective across all types of educational institutions (public, private, and parochial schools), first effective for the 2021/22 school year. Students enrolled in kindergarten through twelfth grade who, on or before the act’s effective date, had already obtained a religious exemption under the prior law, were allowed to retain their exemption. ^21^ Unlike similar policies adopted in other states, Connecticut’s repeal was enacted outside the context of an ongoing outbreak, creating a valuable opportunity to examine the effects of exemption repeal in a non-crisis setting. We therefore aimed to assess whether Connecticut’s repeal of religious vaccine exemptions was associated with changes in kindergarten vaccination coverage and whether post-policy changes differed by school type and county-level religious congregation density. We hypothesized that post-policy increases in vaccination coverage would be greater in private schools, where baseline exemption use has historically been higher,^22^ and in counties with higher religious concentration.

## Methods

We conducted an interrupted time series analysis using state-level data to evaluate changes in kindergarten vaccination coverage associated with Connecticut’s 2021 repeal of religious vaccine exemptions. Outcomes included annual kindergarten vaccination coverage for measles-mumps-rubella (MMR), diphtheria-tetanus-acellular pertussis (DTaP), inactivated polio (IPV), varicella, and hepatitis B.

We obtained annual kindergarten enrollment and vaccination coverage from the Connecticut Department of Public Health. Data were extracted separately for public and private schools and covered school years 2012/13 through 2024/25.^23^ The mean enrollment per school and estimated number of kindergarten-eligible children were compared between the baseline and final study years. Kindergarten-eligible cohorts were approximated from Connecticut birth counts based on the state’s eligibility cutoff, which requires children to be five years old on or before September 1 of the school year to enroll in kindergarten.

We used segmented regressions within an interrupted time series framework to model annual vaccination coverage as a function of time, treating school year 2021/22 as the first post-policy year. Models were parameterized with a continuous time variable, an indicator for the post-policy period, and a time-since-intervention term. Using generalized linear models with a Gaussian distribution and identity link, we modeled vaccine coverage to interpret effects in percentage-point units. Residual autocorrelation was assessed, and Newey-West standard errors were used to account for autocorrelation and heteroskedasticity in the annual time series. We fit separate models for each vaccine overall and stratified by public and private schools.

To examine whether local religious context modified policy response, we used county-level religious congregation density, defined as congregations per 10,000 residents, as a proxy for local religious context. Data were extracted from the Internal Revenue Service National Center for Charitable Statistics Business archives.^24^ Counties were categorized as high- or low-religiosity using the median congregation density as the cut point. We then fit analogous interrupted time series models for the two groups and compared pre-policy slopes, immediate level changes, and post-policy slope changes in vaccine coverage.

To contextualize trends, we used Arizona, Louisiana, and Oregon as comparison states in an analogous interrupted time series analysis of MMR vaccination coverage. MMR was selected for this analysis because it is the vaccine most directly linked to exemption-related outbreak risk. Comparison states were selected a priori based on availability of annual kindergarten MMR coverage and religious congregation density data, broad similarity to Connecticut in population size and health insurance coverage,^25^ and retention of religious exemptions throughout the study period, providing a stable policy comparison for Connecticut’s repeal of religious vaccine exemptions.

The overall impact of the policy was estimated as the difference between Connecticut’s MMR coverage in the final study period, and the predicted counterfactual coverage had the pre-policy trend continued in Connecticut. We also compared Connecticut’s MMR coverage in the final study period with observed MMR coverage in the comparison states. Cumulative impact was translated into an estimated number of additional immunized kindergarteners by summing annual enrollment-weighted differences between observed and counterfactual MMR coverage across post-policy years. Analyses were conducted in R version 4.5.1 and Stata, version 19 (StataCorp). Statistical significance was defined as a two-sided p < 0.05. The study used publicly available aggregate data and was not considered human subjects research.

## Results

### Overall Trends in Kindergarten Enrollment and Vaccine Coverage in Connecticut

In the baseline school year, 41,604 kindergarteners were enrolled across 729 schools in Connecticut, including 562 public and 167 private schools. Between the baseline and final study years, the estimated number of kindergarten-eligible children in Connecticut based on birth month declined by 17.7%, while mean enrollment per school declined by 13.2% overall. The decline in public-school enrollment per school was similar in magnitude to the birth-cohort decline, whereas private-school enrollment remained relatively stable (eTable 1). During the pre-policy period, kindergarten vaccination coverage in Connecticut was steadily declining at rates ranging from −0.16% to −0.20% per year (p < 0.001 for all vaccines). Following repeal of religious exemptions, these trends reversed, with all vaccines demonstrating immediate level increases of 0.43% to 0.68% and upward shifts in post-policy trends, resulting in year-over-year increases of 0.88% to 1.02% across vaccines (eTable 2).

### Vaccination Trends by School Type

At baseline, vaccination coverage for all vaccines was higher in public schools than in private schools (Figure 1). Before policy implementation, coverage declined significantly in both public and private schools, but the magnitude of decline did not differ significantly between the two settings (eTable 3). Following policy implementation, coverage increased in both groups, but post-policy gains were larger in private schools, with significant between-group differences for varicella (private: 1.65% vs public: 1.02%, p = 0.002), DTaP (private: 1.67% vs public: 0.87%, p = 0.007), polio (private: 1.68% vs public: 0.90%, p = 0.004), and hepatitis B (private: 2.01% vs public: 0.85%, p < 0.001).

**Figure 1.**
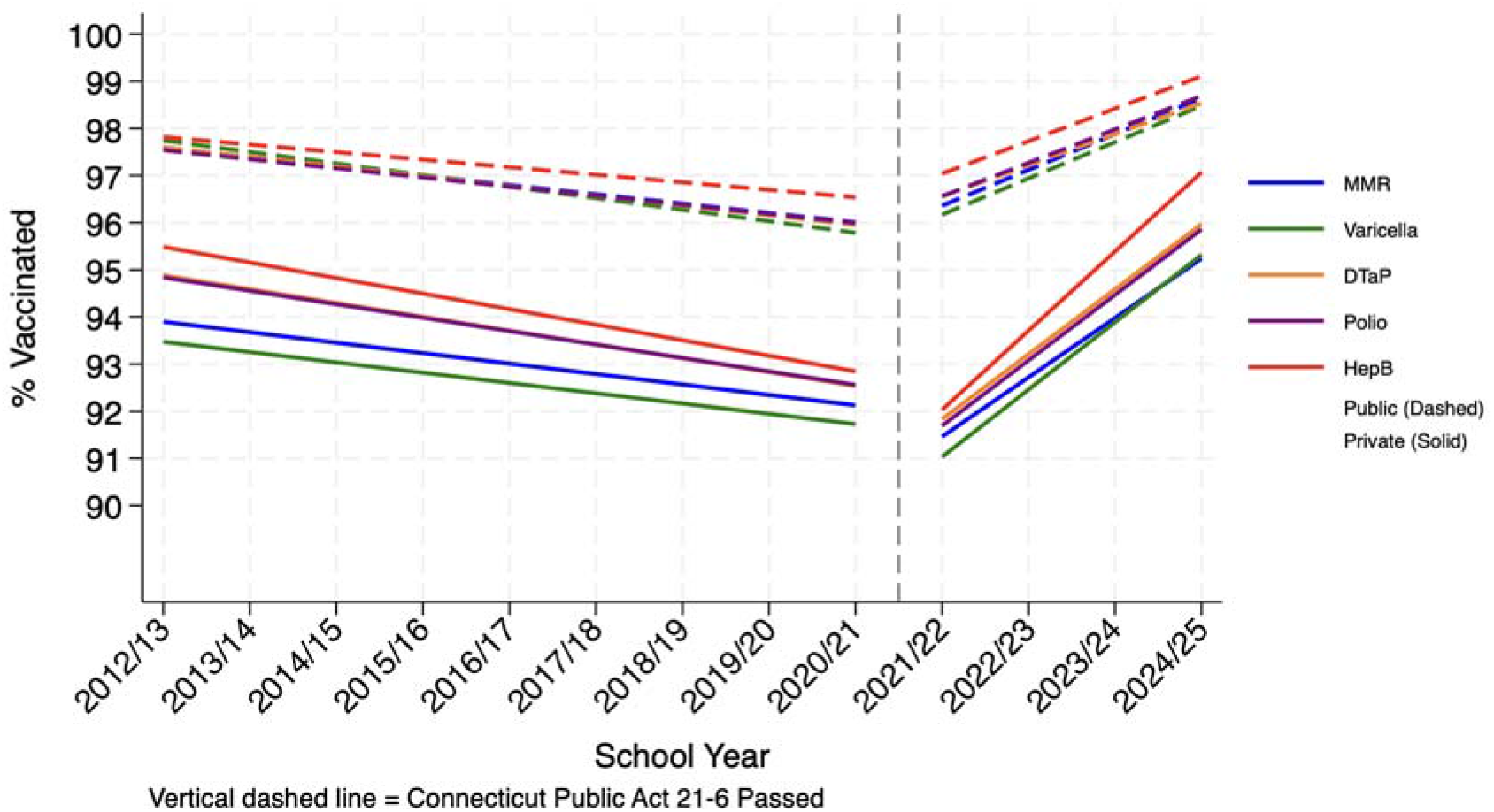
Kindergarten Vaccination Coverage in Connecticut by Vaccine and School Type Before and After Religious Exemption Repeal, 2012-2025.

### Vaccination Trends by Religiosity

At baseline, Connecticut had 4.286 religious congregations per 10,000 inhabitants, the most common religious groups being Catholic, Protestant, and Muslim. By 2024, congregation density increased to 5.510 per 10,000 population. At the state level, baseline religious congregation density differed between Connecticut and the comparison states, although pre-policy religious density trends were similar across states (eTable 4).

At the county-level, vaccination coverage declined before policy implementation in both high- and low-religiosity counties, with similar pre-policy slopes (high: −0.125% per year vs low: −0.189% per year, p = 0.268) (eFigure 1). After implementation of PA 21-6, coverage across all vaccines increased at a similar rate in both high- and low-religiosity counties (eTable 5).

### Vaccination Trends Relative to Other States

At baseline, kindergarten MMR vaccination coverage was 97.1% in Connecticut, 94.5% in Arizona, 93.5% in Oregon, and 96.6% in Louisiana. Pre-intervention coverage declined across all states, with no significant differences in trends between Connecticut and the comparison states. After policy implementation, MMR coverage differed significantly between Connecticut and all comparison states. Specifically, coverage increased in Connecticut, reversing the pre-intervention decline, while it continued to decline in Arizona, Oregon, and Louisiana (Table 1). By the 2024/25 school year, Connecticut’s MMR coverage had increased to 98.2%, which exceeded that of all comparison states by 5.6% to 9.6% (Figure 2). Under the counterfactual scenario, which modeled expected coverage in Connecticut had the policy not been implemented, coverage at the last study period would have been 95.0%, a difference of 3.21% (95% CI, 2.53 to 3.90). Cumulatively, the model-estimated policy effect corresponded to 2,579 additional kindergarteners immunized against MMR compared with the no-policy counterfactual (eTable 6).

**Table 1.**
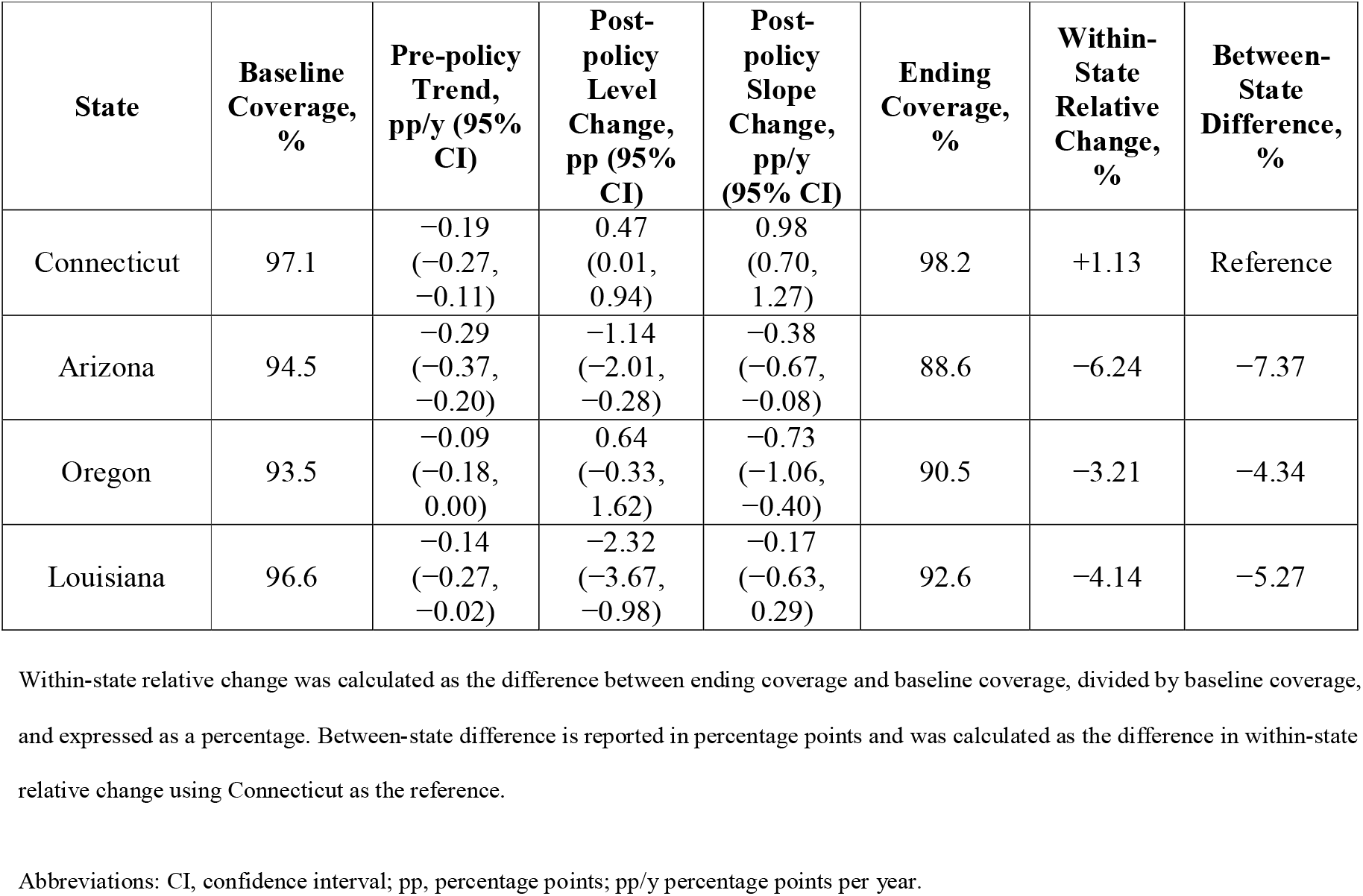
Kindergarten MMR Vaccination Coverage and Policy-Associated Changes in Connecticut Relative to Comparison States.

**Figure 2.**
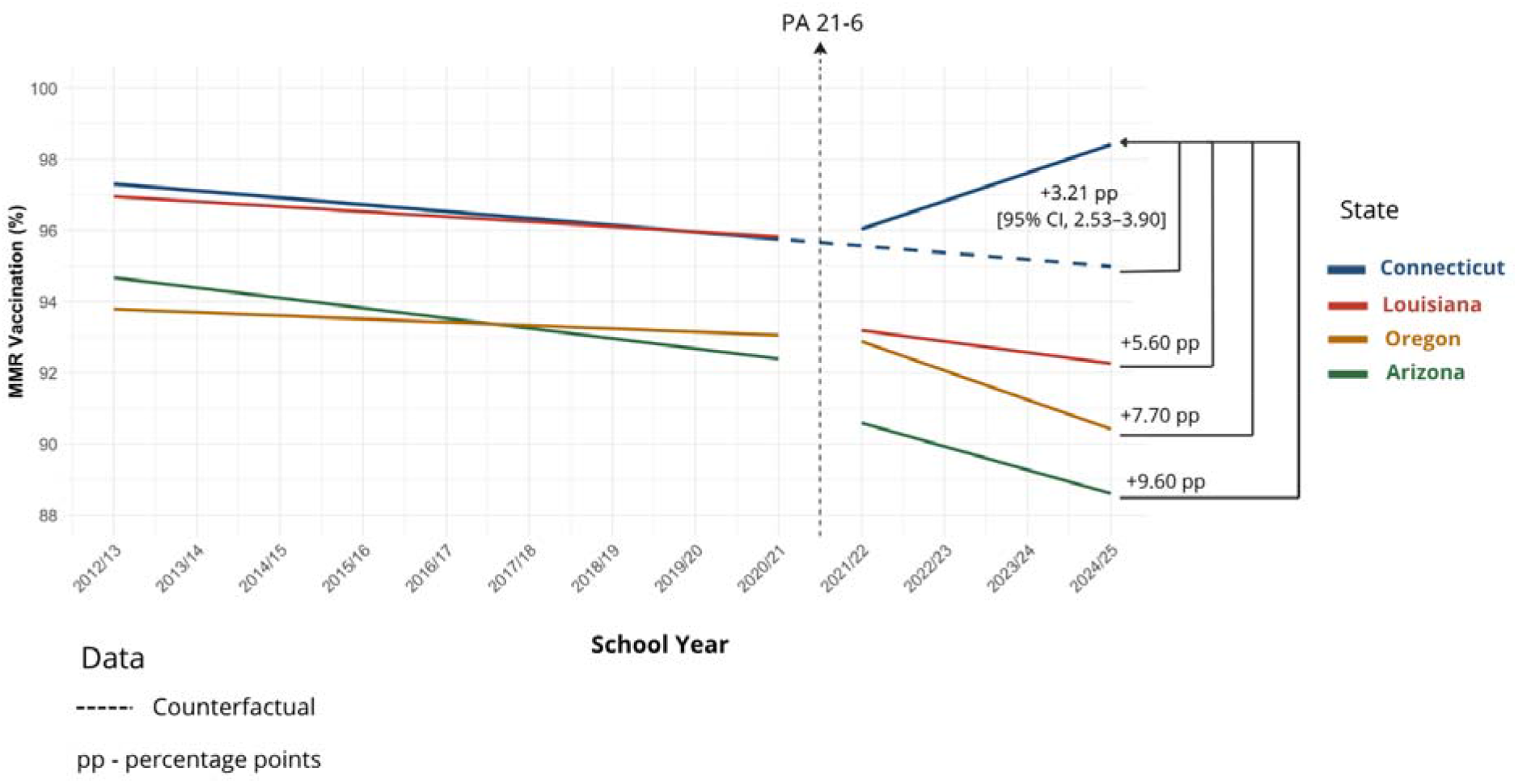
Interrupted time-series analysis of MMR vaccination coverage in Connecticut and comparison states, 2012-2025.

## Discussion

Connecticut’s 2021 repeal of religious exemptions to school vaccination requirements was associated with a clear reversal of previously declining kindergarten vaccination coverage across all studied vaccines. Post-policy gains were observed statewide, but were larger in private than public schools, suggesting narrowing of pre-existing differences in school-entry vaccination coverage. Notably, these gains occurred despite increasing religious congregation density over the study period. The divergent trajectories observed in Connecticut and the comparison states strengthen the interpretation that the post-policy increase in coverage was not simply attributable to broader secular trends in vaccination uptake.

A growing body of state-level evidence suggests that exemption reform can improve school-entry vaccination coverage. In California, repeal of nonmedical exemptions was associated with reduced potential for within-school contact among under-immunized kindergarteners and no corresponding increase in homeschooling rates.^17–20^ In New York, elimination of religious exemptions led to substantial declines in exemption rates and significant increases in vaccination coverage, while in Washington, repeal of the personal belief exemption for MMR was followed by a 5.4% relative increase in kindergarten MMR completion.^26^ Our study extends this evidence in an important way because Connecticut’s repeal of religious exemptions occurred outside the context of an active outbreak, reducing the possibility that post-policy gains were driven primarily by outbreak-related fear, media attention, or intensified public health response.

Nationally, vaccine exemption rates among kindergarteners have continued to rise, reaching 3.3% in the 2023/24 school year.^27^ Private schools in the United States have historically had higher vaccine exemption rates than public schools,^22,27^ likely reflecting clustering of families with similar social, cultural, and ideological beliefs.^12^ This pattern has important public health implications because pockets of under-immunized children can amplify transmission during outbreaks.^8,10^ Recent measles outbreaks have underscored this vulnerability, with most cases in the 2025 multistate outbreak linked to transmission in close-knit communities with low measles vaccination coverage.^28,29^ Our analysis shows that the repeal of religious exemptions reversed prior declines in vaccination coverage across both public and private schools, with larger post-policy gains in private schools. These data suggest that such policies may be particularly effective in settings with historically lower baseline coverage and higher exemption burden, helping to narrow long-standing disparities in school-entry vaccination coverage.

While exemption repeals are often debated in the context of religious freedom, evidence from the largest U.S. faith traditions indicates that routine childhood vaccination is generally permitted and often encouraged.^13^ Consistent with this, the American Academy of Pediatrics notes that no major religious tradition explicitly prohibits vaccination.^30^ For example, the Catholic Church affirms that receiving recommended vaccines is morally acceptable and protects vulnerable populations,^14,31^ and the Greek Orthodox Archdiocese has stated that its tradition provides no doctrinal basis for exemptions from vaccination.^32^ This supports the notion that religious exemptions are not always claimed on the basis of formal doctrinal opposition to vaccination. Connecticut provides an informative case study in this regard. Our data show that repeal of the religious exemption occurred during a period of increasing religious congregation density and was associated with improved school-entry vaccination coverage, without meaningful differences by local religious concentration. These findings suggest that the impact of the policy was not simply attributable to secularization in the state.

This study has several limitations. Our analyses relied on aggregate observational data and therefore could not assess individual-level exemption behavior, vaccination decisions, or motivations for claiming exemptions. Although interrupted time series analysis strengthens causal inference, we cannot exclude the possibility that other contemporaneous factors including changes in healthcare utilization and educational systems during the COVID-19 pandemic period influenced school-entry vaccination coverage. We also cannot exclude the possibility that some families may have responded to the policy by exiting the public or private school system, which could inflate observed school-entry coverage. Connecticut does not have a central database with homeschool counts, so quantifying the number of children who exited public or private schools was not possible for this analysis. Notably, prior work from California after repeal of nonmedical exemptions did not find a corresponding increase in homeschooling rates.^20^ In addition, our religiosity analysis used county-level religious congregation density as a proxy for local religious context, which may not fully capture individual belief or doctrinal commitment.

## Conclusion

Connecticut’s 2021 repeal of religious exemptions was associated with consistent and sustained improvements in childhood vaccination coverage, reversing prior declines across multiple vaccines. Increases were observed in both public and private schools and did not differ significantly according to county-level religious congregation density. Taken together, these findings suggest that repeal of religious exemptions may improve school-entry vaccination coverage even during a period of increasing religious congregation density and may help reduce pre-existing disparities between public and private schools.

## Data Availability

All data produced in the present study are available upon reasonable request to the authors

