## Supplemental Materials for "Impact of Connecticut’s 2021 Repeal of Religious Vaccine Exemptions on Kindergarten Vaccine Coverage"

**eTable 1. Changes in Kindergarten-Eligible Children and Mean Enrollment per School**

| **Measure** | **School Year**  **2012/13** | **School Year**  **2024/25** | **Absolute Change** | **Relative Change (%)** |
| --- | --- | --- | --- | --- |
| Estimated birth cohort | 42,064 | 34,622 | −7,442 | −17.69 |
| Kindergarten enrollment | 41,604 | 30,112 | −11,492 | −27.62 |
| All schools, mean (n/N) | 57.07 (41,604/729) | 49.53 (30,112/608) | −7.54 | −13.21 |
| Public schools, mean (n/N) | 67.61 (37,999/562) | 55.28 (27,973/506) | −12.33 | −18.24 |
| Private schools, mean (n/N) | 21.59 (3,605/167) | 20.97 (2,139/102) | −0.62 | −2.87 |

2012/13 kindergarten cohort: Connecticut births from September 2006 through August 2007; 2024/25 kindergarten cohort: Connecticut births from September 2018 through August 2019.

**eTable 2. Interrupted Time-Series Estimates for Kindergarten Vaccination Coverage in Connecticut, 2012-2025.**

| **Vaccine** | **Baseline Coverage, % (95% CI)** | **Pre-policy Trend, pp/y (95% CI)** | **Post-policy**  **Level Change,**  **pp (95% CI)** | **Post-policy**  **Slope Change,**  **pp/y (95% CI)** |
| --- | --- | --- | --- | --- |
| MMR | 97.31  (96.94, 97.67)** | −0.19  (−0.27, −0.11)** | 0.47  (0.01, 0.94)* | 0.98  (0.69, 1.27)** |
| Varicella | 97.04  (96.70, 97.38)** | −0.19  (−0.27, −0.11)** | 0.43  (0.01, 0.86)* | 1.02  (0.77, 1.26)** |
| DTaP | 97.37  (97.06, 97.68)** | −0.20  (−0.28, −0.13)** | 0.68  (0.29, 1.08)** | 0.89  (0.68, 1.11)** |
| Polio | 97.39  (97.04, 97.74)** | −0.20  (−0.29, −0.11)** | 0.68  (0.22, 1.13)** | 0.94  (0.71, 1.18)** |
| HepB | 97.64  (97.29, 97.98)** | −0.16  (−0.22, −0.10)** | 0.59  (0.20, 0.98)** | 0.88  (0.64, 1.12)** |

Baseline coverage represents estimated kindergarten vaccination coverage during the 2012/13 school year. Pre-policy and post-policy slope estimates reflect annual percentage-point changes in vaccination coverage before and after implementation of Connecticut Public Act 21-6 (2021/22 school year), while level-change estimates represent the immediate change in coverage following policy implementation.

Abbreviations: CI, confidence interval; pp, percentage points; pp/y percentage points per year; MMR, measles-mumps-rubella; DTaP, diphtheria-tetanus-acellular pertussis; HepB, hepatitis B. Statistical significance: **p*<0.05; **p<0.01.

**eTable 3. Interrupted Time-Series Estimates for Kindergarten Vaccination Coverage by Vaccine and School Type.**

| **Vaccine** | **School Type** | **Baseline Coverage, % (95% CI)** | **Pre-policy Trend, pp/y (95% CI)** | **Post-policy Level Change, pp (95% CI)** | **Post-policy Slope Change, pp/y**  **(95% CI)** | **Difference in Post-policy Trend, pp/y**  **(95% CI)** |
| --- | --- | --- | --- | --- | --- | --- |
| MMR | Public | 97.59 (97.16, 98.03)** | −0.20 (−0.30, −0.09)** | 0.55 (−0.002, 1.11) | 0.96 (0.63, 1.28)** | 0.50 (-0.04, 1.04) |
|  | Private | 93.90 (93.16, 94.64)** | −0.22 (−0.41, −0.03)* | -0.44 (-2.45, 1.56) | 1.48 (1.06, 1.90)** |  |
| Varicella | Public | 97.75 (96.74, 98.76)** | −0.24 (−0.42, −0.07)** | 0.63 (0.02, 1.23)* | 1.02 (0.73, 1.30)** | 0.66 (0.24, 1.08)** |
|  | Private | 93.47 (92.77, 94.18)** | −0.22 (−0.39, −0.05)* | -0.48 (-2.12, 1.17) | 1.65 (1.31, 1.99)** |  |
| DTaP | Public | 97.60 (97.24, 97.97)** | −0.21 (−0.30, −0.11)** | 0.82 (0.35, 1.29)** | 0.87 (0.69, 1.04)** | 0.72 (0.20, 1.24)** |
|  | Private | 94.88 (94.47, 95.30)** | −0.29 (−0.40, −0.19)** | -0.41 (-1.94, 1.11) | 1.67 (1.20, 2.15)** |  |
| Polio | Public | 97.54 (97.17, 97.91)** | −0.19 (−0.29, −0.10)** | 0.76 (0.27, 1.25)** | 0.90 (0.72, 1.08)** | 0.68 (0.21, 1.15)** |
|  | Private | 94.84 (94.44, 95.25)** | −0.29 (−0.38, −0.19)** | -0.58 (-2.03, 0.86) | 1.68 (1.26, 2.09)** |  |
| HepB | Public | 97.82 (97.45, 98.19)** | −0.16 (−0.23, −0.09)** | 0.66 (0.27, 1.05)** | 0.85 (0.62, 1.08)** | 0.99 (0.44, 1.54)** |
|  | Private | 95.49 (95.01, 95.96)** | −0.33 (−0.46, −0.20)** | -0.47 (-2.16, 1.19) | 2.01 (1.54, 2.48)** |  |

Baseline coverage represents estimated kindergarten vaccination coverage during the 2012/13 school year. Pre-policy and post-policy slope estimates reflect annual percentage-point changes in vaccination coverage before and after implementation of Connecticut Public Act 21-6 (2021/22 school year), while level-change estimates represent the immediate change in coverage following policy implementation. The final column represents the differential change in vaccination trends between private and public schools (private − public), corresponding to the interrupted time series interaction term.

Abbreviations: CI, confidence interval; pp, percentage points; pp/y percentage points per year; MMR, measles-mumps-rubella; DTaP, diphtheria-tetanus-acellular pertussis; HepB, hepatitis B. Statistical significance: **p*<0.05; **p<0.01.

**eTable 4: Baseline Characteristics and Trajectories of Religious Congregation Density (2012–2024)**

| **State** | Baseline Congregation Density (2012) | Annual Pre-Policy Trends (β, 95% CI) ᵃ | Post-2021 Policy Divergence (β, 95% CI)ᵇ |
| --- | --- | --- | --- |
| Connecticut | 4.29 | 0.11 (0.06, 0.17)** | 0.37 (−0.54, 1.28) |
| Arizona | 4.83 | 0.05 (0.01, 0.08)* |  |
| Oregon | 6.53 | 0.02 (−0.02, 0.06) |  |
| Louisiana | 4.79 | 0.12 (0.09, 0.16)** |  |

Density is measured as the number of religious congregations per 10,000 residents.

ᵃ Omnibus parallel trends test evaluating the interaction between time and treatment status during the pre-policy window (2012-2020) confirmed that the annual trajectories were statistically similar and parallel across states (β interaction = 0.05, SE = 0.10, p = 0.61).

ᵇ Computed via a trend-adjusted difference-in-differences interaction model (p = 0.42), evaluating Connecticut's post-2021 trajectory deviation against the pooled comparison states.

Abbreviations: CI, confidence interval; β, regression coefficient. Statistical significance: **p*<0.05; **p<0.01.

**eTable 5. Interrupted Time-Series Estimates for Kindergarten Vaccination Coverage by County-Level Religious Congregation Density**

| **Group** | **Baseline Coverage (95% CI)** | **Pre-policy Trend, pp/y (95% CI)** | **Post-policy Level Change, pp (95% CI)** | **Post-policy Slope Change, pp/y (95% CI)** | **Difference in Post-policy Trend, pp/y (95% CI)** |
| --- | --- | --- | --- | --- | --- |
| High-religiosity counties | 97.11 (96.74, 97.48) | −0.13 (−0.20, −0.05) | −0.88 (−1.90, 0.15) | 1.03 (0.57, 1.48) | 0.34 (−0.27, 0.95) |
| Low-religiosity counties | 97.02 (96.59, 97.45) | −0.19 (−0.27, −0.11) | −1.13 (−2.51, 0.25) | 1.36 (0.73, 2.00) |  |

Counties were classified as high or low religiosity based on median pre-policy (2012–2020) religious congregation density per 10,000 population. New London, New Haven, Hartford, and Fairfield counties were categorized as high religiosity, whereas Tolland, Windham, Litchfield, and Middlesex counties were categorized as low religiosity; classifications remained stable across study years.

Abbreviations: CI, confidence interval; pp, percentage points; pp/y percentage points per year.

**eTable 6. Observed vs Counterfactual MMR Vaccination Counts Among Connecticut Kindergarteners After Policy Implementation**

| School Year | Enrolled Kindergarteners | Kindergarteners Vaccinated Against MMR | Estimated Kindergarteners Vaccinated Against MMR in Counterfactual Scenario |
| --- | --- | --- | --- |
| 2021/22 | 35,451 | 33,913 | 33,891 |
| 2022/23 | 35,580 | 34,633 | 33,943 |
| 2023/24 | 36,184 | 35,354 | 34,447 |
| 2024/25 | 30,112 | 29,566 | 28,606 |
| Total: | | 133,466 | 130,887 |
| Difference: | | 2,579 | |

Counterfactual estimates represent the projected number of Connecticut kindergarteners vaccinated against MMR had pre-policy vaccination trends continued without implementation of Public Act 21-6. The difference represents the cumulative excess number of kindergarteners vaccinated against MMR during post-policy school years relative to the modeled counterfactual scenario.

Abbreviations: MMR, measles-mumps-rubella.

**eFigure 1. Kindergarten Vaccination Coverage in Connecticut by County-Level Religious Congregation Density Before and After Religious Exemption Repeal.**

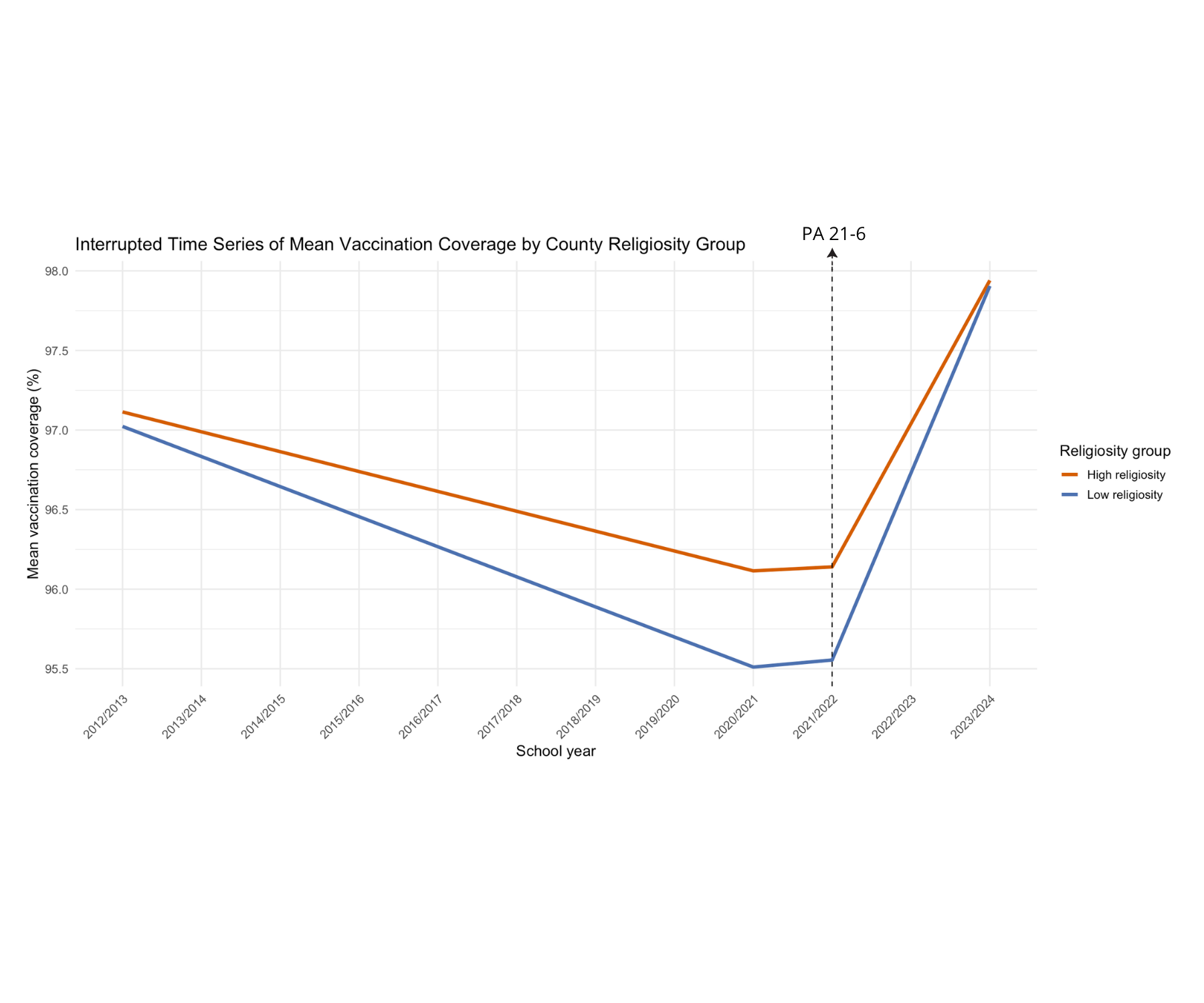
